# Rapid development and field evaluation of a portable CRISPR-based assay for Mpox during the 2025 Sierra Leone outbreak

**DOI:** 10.1101/2025.10.08.25337506

**Authors:** Nisha Gopal, Tsion Abay, Carolyn Payne, Michael Gomez, Maariam Manjia Rogers, Ibrahim Umaru Fofanah, Tiangay PMS Kallon, Mohamed S. Kamara, Ho-Jun Suk, John Demby Sandi, Taylor Brock-Fisher, Elyse Stachler, Lao-Tzu Allan-Blitz, David J. Roach, Marietou F Paye, Colby Wilkason, Donald S. Grant, Al Ozonoff, Pardis C. Sabeti

**Author notes:** These authors jointly supervised the work (Al Ozonoff, Pardis C. Sabeti).

## Abstract

In early 2025, Sierra Leone experienced a large outbreak of mpox clade IIb, underscoring the urgent need for portable, low-cost diagnostics in decentralized settings. We rapidly developed and field-deployed Mpox SHINE, a CRISPR–Cas13 assay that integrates lyophilized reagents, ambient-temperature lysis, and automated fluorescence detection on a portable device, the DxHub. The assay achieved analytical sensitivity down to 10 copies/µL with minimal hands-on time. In-country evaluation of 56 clinical specimens showed complete concordance with qPCR, with 100% sensitivity (45/45) and 100% specificity (11/11) at the sample level. Mpox SHINE also detected the virus directly from unextracted lesion swabs, maintaining 100% sensitivity and specificity in 16 samples (8 positive, 8 negative). Across extracted and unextracted samples, the mean time-to-result was ∼35 minutes, with all positives detected within 45 minutes. Thus, Mpox SHINE performed on the DxHub demonstrates how CRISPR-based pathogen detection can be rapidly translated into portable tools for the front lines of outbreak response.

**Teaser:** CRISPR-Cas13 assay and portable device bring rapid, reliable mpox testing to outbreak settings.

## Introduction

In early 2025, Sierra Leone faced a fast-moving outbreak of mpox clade IIb that infected thousands and strained the country’s public health system. By July, more than 4,000 confirmed cases had been reported, demonstrating both the scale of the crisis and the risk of wider regional spread (*1–5*).

Mpox is caused by the mpox virus (MPXV), an enveloped double-stranded DNA Orthopoxvirus in the Poxviridae family, and spreads through both animal-to-human and human-to-human transmission, often via close contact with lesions and other bodily fluids (*6*). This crisis followed on the heels of successive mpox resurgences worldwide: a global multi-country outbreak of clade IIb in 2022, and a surge of clade I, including lineage Ib, across central Africa in 2024, when the World Health Organization (WHO) declared mpox a public health emergency of international concern (*7*, *8*).

Despite the escalating global health threat of mpox, diagnostics remain constrained worldwide (*9*). Testing depends almost entirely on qPCR, or expensive cartridge-based systems (*10*), performed in a handful of centralized laboratories. Although qPCR is highly sensitive, it requires costly infrastructure, specialized equipment, and trained personnel, limiting accessibility in rural districts experiencing the heaviest caseloads. As a result, frontline clinics are left without access to timely results, a problem exacerbated by severe testing shortages that create backlogs and multi-day delays at the very moment when rapid detection is most critical. Unlike other viral diseases, antibody-based assays for mpox are not widely available nor are any clinically approved point-of-care (POC) tests for mpox available for distribution at decentralized sites throughout Sierra Leone. The result is a widening diagnostic gap: community surveillance stalls, delayed sample turnaround time, and the outbreak spreads faster than the system can respond.

CRISPR-based nucleic acid detection strategies provide a promising solution to POC diagnostic gaps (*11*). By combining isothermal amplification with programmable Cas enzymes, they can achieve sensitivity and specificity comparable to qPCR while enabling portable, extraction-free formats suitable for field use (*12*, *13*). Artificial Intelligence (AI)-guided assay design makes it possible to generate new CRISPR tests within weeks (*14*), while lyophilized reagents, ambient-temperature lysis, and simplified workflows reduce the need for equipment or cold chain, allowing assays to be rapidly distributed and performed at the point of need. Despite recent progress, the broader potential of CRISPR diagnostics remains underutilized, highlighting the need for platforms that can rapidly translate design innovations into routine deployment at the front lines.

Here we introduce Mpox SHINE (Streamlined Highlighting of Infections to Navigate Epidemics) (*15*), developed and deployed via the DxHub platform (DxLab Inc., Somerville, MA, USA) in direct response to the Sierra Leone clade IIb outbreak. In response to the outbreak, we advanced from assay design to on-site validation in Sierra Leone within a single development cycle—encompassing primer and crRNA design, lyophilization, device integration, and clinical evaluation at Kenema Government Hospital. The sections that follow describe assay design, analytical performance on synthetic targets, device integration, and evaluation on both extracted and minimally processed clinical samples.

## Results

### Design, optimization, and validation of a SHINE assay for Mpox Clade IIb

To rapidly respond to the 2025 Sierra Leone mpox outbreak, we developed a CRISPR-Cas13 assay using the SHINE framework, which combines recombinase polymerase amplification (RPA) with Cas13 detection in a single tube and incorporates AI-guided design via ADAPT (*14*, *15*).

Using an AI-guided SHINE approach, we identified a highly conserved amplicon as the optimal target for mpox clade IIb detection. From 33 newly sequenced genomes generated at Kenema Government Hospital in April–May 2025 (*16*); **Supplementary Table 3**), we designed forward and reverse RPA primers and a Cas13 crRNA with complete coverage across these sequences **(Supplementary Tables 1, 3)**. The ADAPT-derived amplicon was an 88 bp segment in the 3′ variable region between gp161 and gp162, upstream of the inverted terminal repeat (NC_063383) **(Figure 1a, Supplementary Table 1)**. This locus is fully conserved across clade IIb and highly conserved in clade IIa (97.6%) but absent from clade I (Ia/Ib) due to deletions seen by gaps in our sequence alignment. In comparison, sequence identity to other Orthopoxviruses was lower (vaccinia 91.3%, cowpox 91.3%, camelpox 86.3%), reflecting divergence from non-mpox orthopoxes while retaining broad coverage of outbreak-relevant lineages.

**Figure 1.**
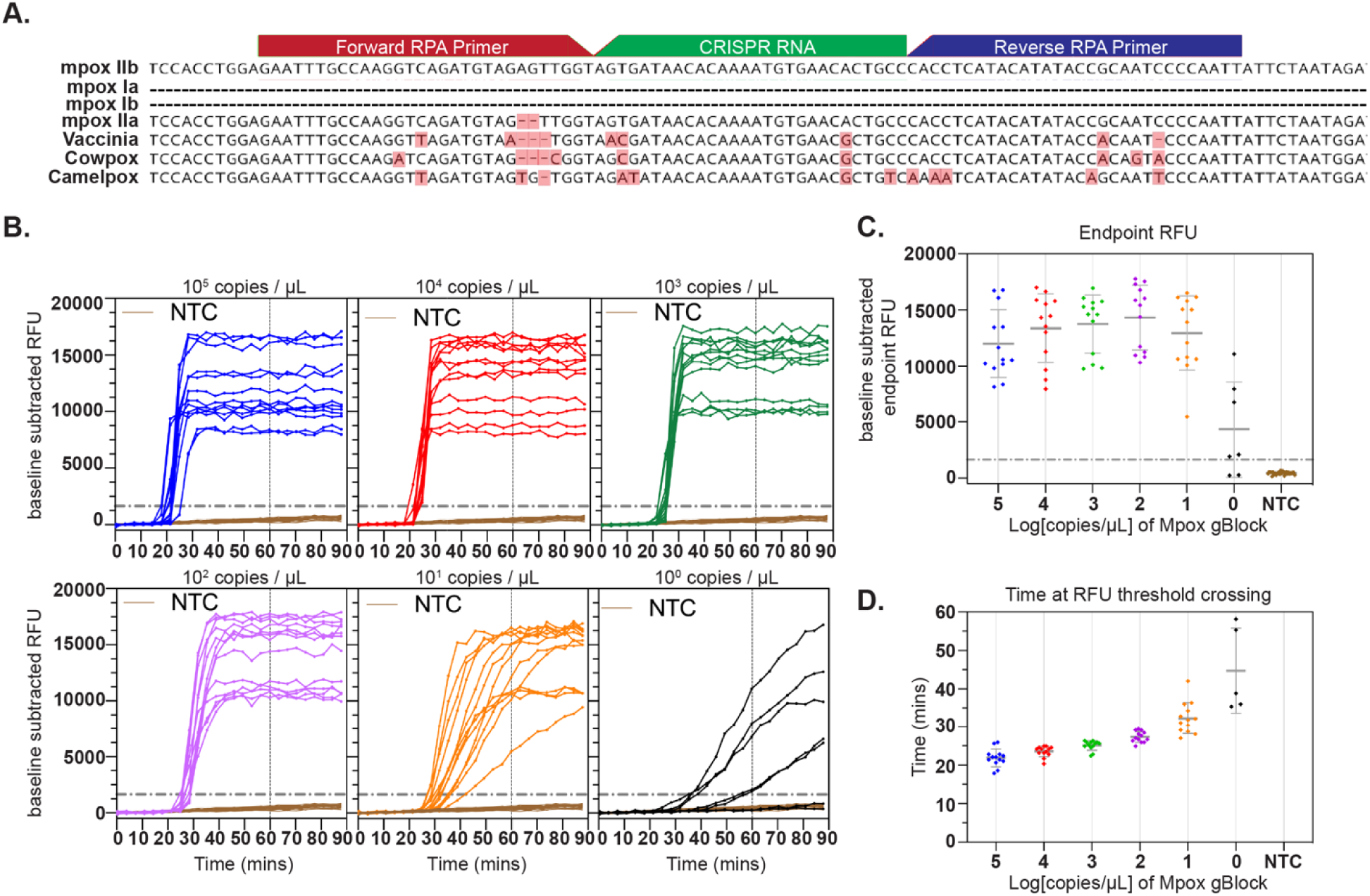
Mpox SHINE assay design and analytical performance on a plate reader. **(A)** Schematic of the Mpox target locus. Forward and reverse RPA primers (red/blue) and the crRNA spacer (green) are shown aligned to the MPXV clade IIb genome (“Pathoplexus | Mpox Virus Genomes,” n.d.) together with MPXV clade IIa (97.5% identity; “Pathoplexus | Mpox Virus Genomes,” n.d.) and representative Orthopoxvirus genomes: vaccinia (NC_006998.1, 91.3% identity), cowpox (NC_003663.2, 91.3% identity), and camelpox (AY009089.1, 86.3% identity). The amplicon lies in a conserved region of clade II Mpox, with primer and guide sequences listed in **Supplementary Table 1. (B)** Plate reader measured fluorescence kinetics (baseline-subtracted RFU; baseline = mean RFU from 1–4 min) for a 10-fold dilution series of gBlock (10^5^ to 10^0^ copies /µL of reaction) and NTCs. Solid lines indicate individual replicates of Mpox SHINE reactions, and the dotted horizontal line indicates the fixed RFU threshold (NTC mean at 60 min + 10 SD) used for result calling. **(C)** Endpoint RFU at 60 min for the same datasets as **(B)**, plotted as individual technical replicates (pooled from two experiments) for 10^1^–10^5^ copies/µL (n = 13), 10^0^ copies/µL (n=7), and NTCs (n = 32). Horizontal bars denote the mean and error bars denote the standard deviation. The dotted horizontal line indicates the fixed RFU threshold (NTC mean at 60 min + 10 SD). **(D)** Time to RFU threshold crossing for the same datasets, with all replicates corresponding to the same datasets in panel B-C. Horizontal bars denote the mean and error bars denote the standard deviation.

We optimized the Mpox SHINE assay to maximize performance and sensitivity using a synthetic double stranded DNA target containing the selected amplicon **(Supplementary Table 1)**. We tuned primer concentration, primer composition, and magnesium concentration systematically to improve signal (**Supplementary Figure 1**). We performed reactions with lyophilized SHINE reagents containing the newly designed Mpox primers and crRNA **(Figure 1a)**. To assess performance across a dynamic range, we tested a tenfold dilution series of gBlock standards from 10^5^ to 10^0^ copies/µL at 38 °C. We monitored fluorescence on a Biotek Cytation 5 plate reader every 20 seconds for at least 60 minutes **(Figure 1b, c)**.

The optimized assay achieved highly sensitive detection, with a limit of detection (LOD) between ∼1 and ∼10 copies/µL. Using a fixed fluorescence threshold (1644.9 RFU) defined as the mean of no template controls (NTC) plus 10 standard deviations (SD), we detected ∼1 copy/µL in 5 of 7 replicates (71.4%, 95 % CI 35.9–91.8) and ∼10 copies/µL in 13 of 13 replicates (100 %, 95 % CI 77.2–100.0). These results show that the assay consistently reached single digit copy sensitivity under laboratory conditions.

We next asked whether the assay could also deliver rapid time to detection suitable for field use. Time to detection was defined as the point at which fluorescence signal crossed the fixed threshold, and it was inversely related to input copy number **(Figure 1d)**. At high input levels (10 –10^2^ copies/µL), all replicates crossed the threshold within approximately 21-28 minutes, while the lowest-input samples near the detection limit (10¹–10 copies/µL) were detected within 27–45 minutes. No amplification was observed in NTCs over the 60 min assay window. These results highlight the assay’s speed and reliability in generating clinically actionable results in under one hour.

### Integration of Mpox SHINE on the DxHub portable isothermal device

With the Mpox SHINE assay benchmarked on a plate reader, we next integrated it onto a portable isothermal device to evaluate performance in a POC setting. The DxHub (DxLab Inc., Somerville, MA) is a compact instrument that provides isothermal incubation and dual channel real time fluorescence detection **(Figure 2a)**. Achieving a quantifiable fluorescence readout in real time is a key feature of SHINE, yet this capability is difficult to replicate outside of laboratory plate readers. In prior SHINE studies, POC-intended detection has often relied on lateral flow strips, which provide a simple visual readout but are non-quantitative and less amenable to real-time kinetics (*15*, *17–19*). By contrast, the DxHub enables quantitative fluorescence detection with a simple three-step workflow that required fewer than five minutes of hands-on time (**Figure 2a**).

**Figure 2.**
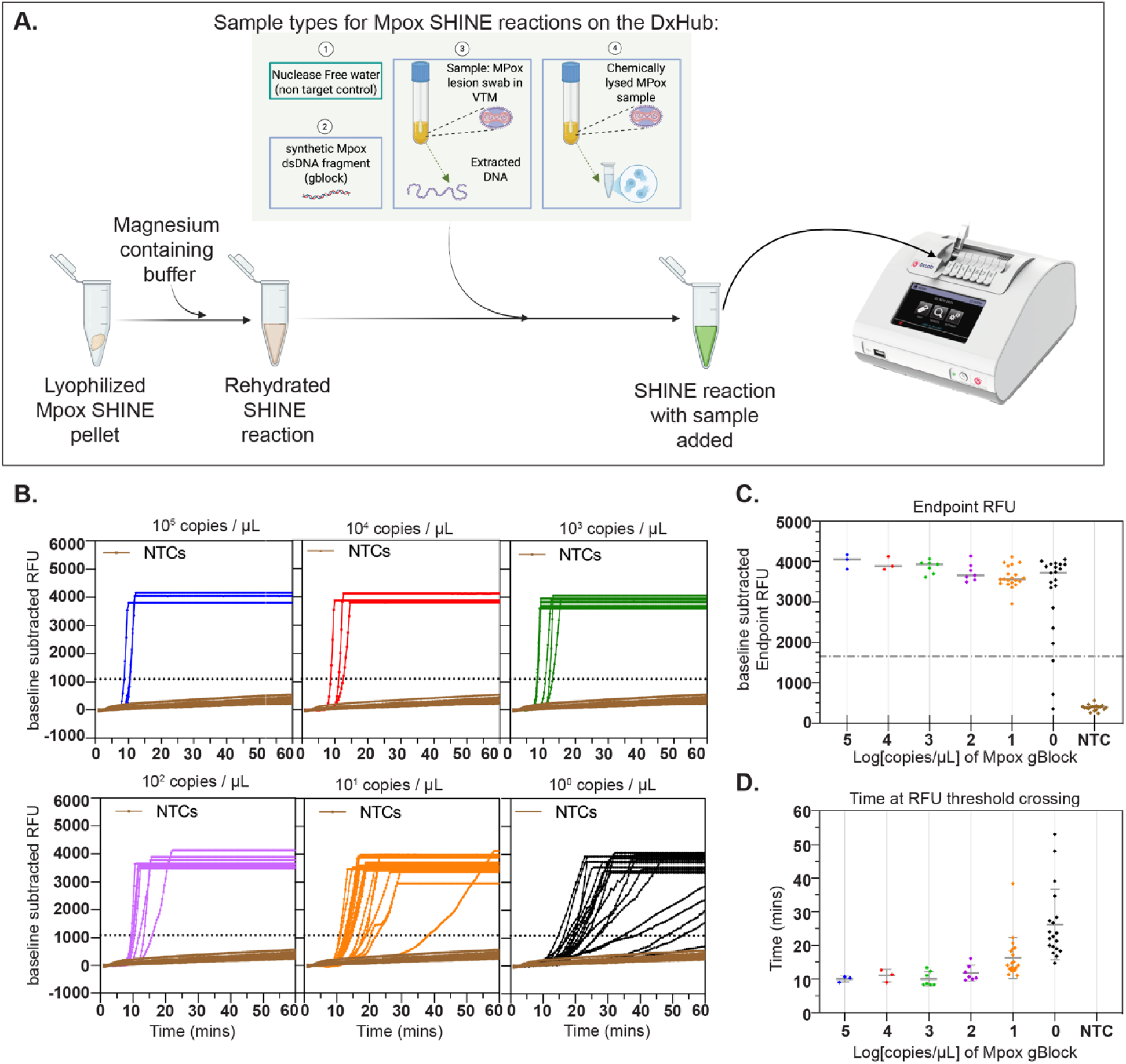
Device integration, workflow, and on-device analytical performance. **(A)** Workflow for running Mpox SHINE on the DxHub. A lyophilized single-reaction pellet is rehydrated with magnesium-containing buffer, sample is added (synthetic dsDNA gBlock, water as a no template control, extracted clinical DNA, or chemically lysed mpox specimen), and the tube is loaded into the device for incubation at 38C along with concurrent fluorescent signal acquisition. VTM = viral transport media. **(B)** On-device fluorescence kinetics (baseline-subtracted RFU; baseline = mean RFU from 1–4 min) for a 10-fold dilution series of gBlock (10^5^ to 10^0^ copies /µL of reaction) and NTCs. Solid lines indicate individual replicates of Mpox SHINE reactions at different gBlock concentrations, and the dotted horizontal line indicates the fixed RFU threshold (NTC mean at 60 min + 10 SD) used for result calling. **(C)** Endpoint RFU at 60 min for the same datasets as **(B)**, plotted as individual technical replicates (pooled from 4 experiments). Replicate counts: 10^0^ copies /µL (n=21), 10^1^ copies /µL (n=20), 10^2-3^ copies /µL (n=7), 10^4-5^ copies /µL (n=3), and NTCs (n=19). Horizontal bars denote the mean and error bars denote the standard deviation. The dotted horizontal line indicates the fixed RFU threshold. **(D)** Time to RFU threshold crossing for the same datasets, with all replicates corresponding to the same datasets in panel **B**-**C**. Horizontal bars denote the mean and error bars denote the standard deviation.

We benchmarked Mpox SHINE performance on the DxHub using a serial dilution of mpox gBlock **(Figure 2b)**. Using a fixed fluorescence threshold defined as the mean of NTCs plus 10 SD (1097.5 RFU), we detected ∼1 copy/µL in 19 of 21 replicates (90.5%; 95% CI 71.1–97.3%) and ∼10 copies/µL in 20 of 20 replicates (100.0%; 95% CI 83.9–100.0%). This yielded an LOD between ∼1 and ∼10 copies/µL for this assay configuration (**Figure 2b, c**).

We next evaluated assay time to detection on the DxHub. Time to detection, as with the plate reader, was defined as the point at which fluorescence signal crossed the fixed threshold and showed the same inverse relationship with input copy number **(Figure 2d)**. Reactions just above the LOD were detected in an average of 16.3 ± 6.1 minutes, with higher-input samples detected in under 15 minutes. Compared to the plate reader **(Figure 1d)**, the DxHub assays resulted in earlier threshold crossings, consistent with differences in fluorescence detection between the two instruments. For end users, this may provide a practical advantage: faster apparent times to result while maintaining equivalent sensitivity and specificity across platforms **(Figures 1c, 2c)**.

### Mpox SHINE performance on extracted and minimally processed clinical samples

Having established the analytical performance of Mpox SHINE in controlled settings, we next evaluated its clinical performance on patient samples at Kenema Government Hospital in Sierra Leone. We analyzed 56 lesion swabs collected in viral transport medium from patients and close contacts. Confirmatory qPCR on freshly extracted DNA identified 45 positives (Ct 15–36) and 11 negatives (**Figure 3a, Supplementary Table 2**), which we used as the reference standard. Our goal was to assess sensitivity, specificity, and time to detection on the DxHub. To do so, we designed two complementary experiments: one using extracted DNA from all specimens tested in triplicate to ensure robustness, and one using chemically lysed, unextracted swabs tested in duplicate to probe performance in a simplified POC workflow.

**Figure 3.**
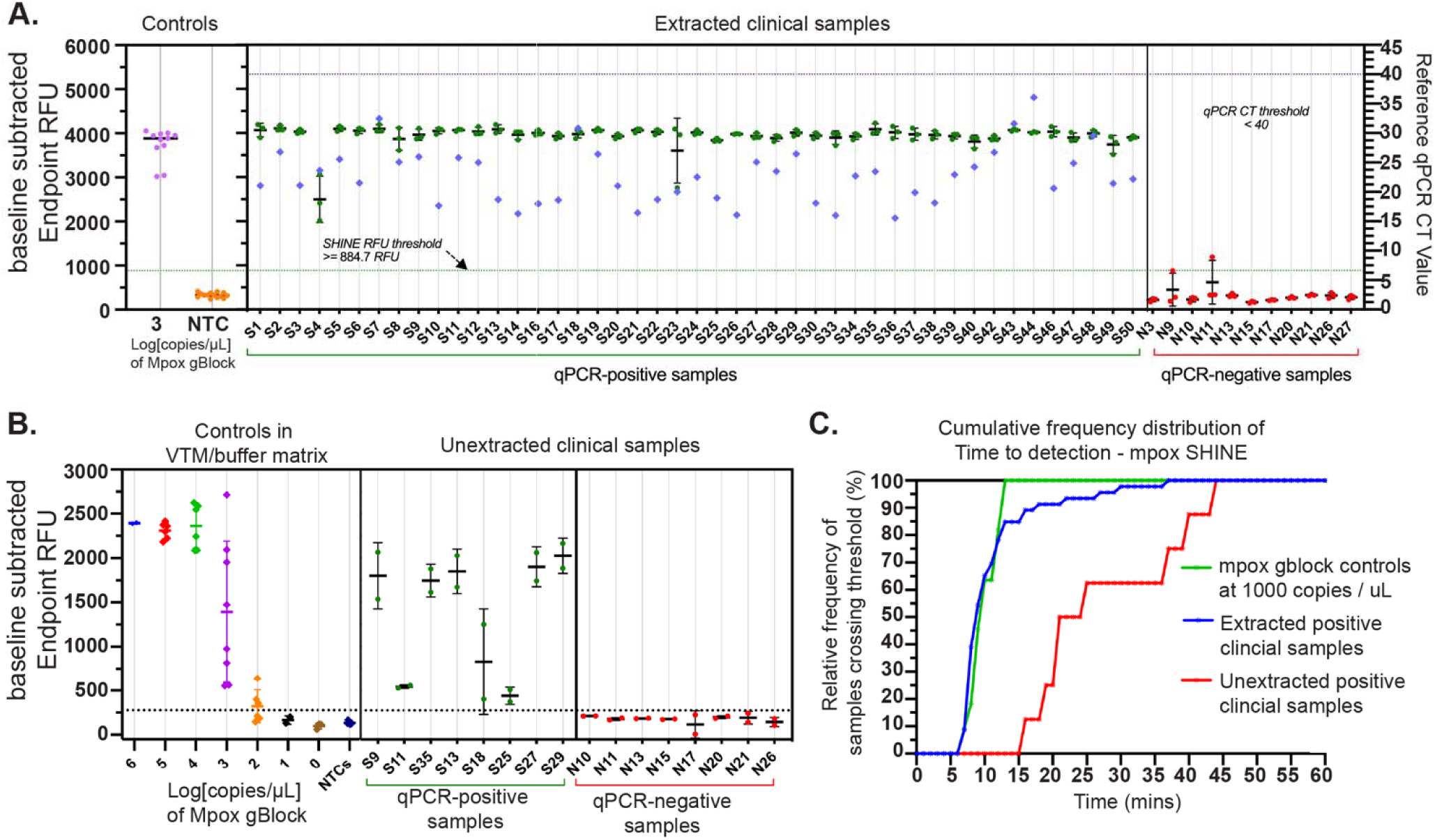
Clinical performance of Mpox SHINE on extracted and unextracted clinical samples. **(A)** Endpoint fluorescence of Mpox SHINE at 60 min (baseline-subtracted RFU; left y-axis) for controls and extracted clinical samples. Control samples are shown in the left panel (Mpox gBlock in lavender [n=11] and NTCs in orange [n=13]). qPCR-positive clinical samples are shown in the middle (green symbols, n=45 positive samples); qPCR-negative clinical samples are shown in the right panel (red symbols, n=11 negative samples). For each Mpox clinical sample, the mean ± SD across 3 technical replicates is shown, along with the corresponding individual replicate values. Blue diamonds indicate the corresponding qPCR Ct value (right y-axis). The dotted green horizontal line marks the fixed RFU threshold based on the mean of NTCs shown on the left side of Panel A, at 60 min + 10 SD. **(B)** Endpoint fluorescence of Mpox SHINE at 60 min (baseline-subtracted RFU; left y-axis) for controls and unextracted clinical samples. Control samples in matrix (VTM + saliva lysis buffer) are shown on the left with a 10-fold dilution series of gBlock from 10^6^ to 10^0^ copies /µL of reaction. Replicate counts: 10^0-1^ copies /µL (n = 2), 10^2^ copies /µL (n =7), 10^3^ copies /µL (n =8), 10^4-5^ copies /µL (n =6), 10^6^ copies /µL (n =2), and NTCs (n=8). qPCR-positive unextracted clinical samples are shown in the middle (green symbols, n=8 positive samples); qPCR-negative unextracted clinical samples are shown in the right panel (red symbols, n=8 negative samples). For each Mpox clinical sample, the mean ± SD across 2 technical replicates is shown, along with the corresponding individual replicate values. The dotted black horizontal line marks the fixed RFU threshold based on the mean of NTCs shown on the left side of Panel B, at 60 min + 10 SD. **(C)** Cumulative frequency distribution of time-to-result for Mpox SHINE on the DxHub. Time-to-detection was defined as the first timepoint at which baseline-subtracted RFU crossed the predefined threshold, and for each sample the mean across replicates was used. Curves show extracted qPCR-positive clinical samples (blue, n=45), unextracted qPCR-positive clinical samples (green, n=8), and 1e3 copies/µL gBlock positive controls (red, n=11).

On extracted samples, Mpox SHINE results closely tracked the qPCR reference standard, yielding a sensitivity of 100% (95% CI 92.1–100.0%) and a specificity of 100% (95% CI 74.1– 100.0%) (**Figure 3a**). All 45 qPCR-positive samples were positive in 3/3 replicates. Of the 11 qPCR-negative samples, 9 were negative in 3/3 replicates, and the remaining 2 were negative in 2 of 3 replicates. In both cases, a single replicate from an otherwise negative sample crossed the fluorescence threshold late in the run, producing low-level amplification that did not replicate across repeat reactions **(Supplementary Figure 2)**. Because SHINE signal kinetics are proportional to input viral load, such weak late signals could represent borderline amplification rather than instrument noise. However, during our assay validation we conducted triplicate testing to enable conservative interpretation by majority rule (2 of 3 replicates), which preserved specificity without masking potential low-level positives. In future diagnostic implementations, similar cases could be resolved through reflex testing, threshold adjustment, or confirmatory retesting workflows.

Together, these results highlight the robustness and interpretability of Mpox SHINE across replicate runs and sample types. Mpox SHINE delivered fast results across extracted clinical samples (**Figure 3c**).

The assay detected positives in an average of 11.4 ± 6.2 minutes, with 85% detected within 15 minutes and 100% within 37 minutes, consistent with the kinetics of positive controls (**Supplementary Figure 5a**). Time to detection tended to increase with higher qPCR Ct values, but the correlation was weak and not statistically significant (Pearson r = 0.20, 95% CI –0.10–0.47, p = 0.1810; Spearman r = 0.46, 95% CI –0.18–0.67, p = 0.0016) (**Supplementary Figure 5b**). Thus, while time to detection broadly mirrored viral load, the assay maintained rapid performance across the full range of Ct values observed.

On unextracted lesion swabs, Mpox SHINE achieved a sensitivity of 100% (95% CI 72.2– 100.0%) and a specificity of 100% (95% CI 72.2–100.0%) relative to qPCR (**Figure 3b**). We tested eight qPCR-positive and eight qPCR-negative samples in duplicate (32 reactions total, **Supplementary Figure 4**). All eight positives were detected, and all eight negatives remained negative. Because matrix-specific controls were not run on-site, thresholds for these analyses were set retrospectively using dedicated controls in the same VTM/buffer background **(Figure 3b**, **Supplementary Figures 3d and 5c)**, informed by prior matrix-development experiments on both the DxHub and a plate reader **(Supplementary Figure 3a-c)**.

Time to detection on unextracted samples was slower than on extracted DNA but remained actionable for outbreak response (**Figure 3c**). The assay identified positives in an average of 27.9 ± 10.7 minutes, with 50% detected within 21 minutes and 100% within 44 minutes, consistent with control reactions in the same VTM/buffer matrix (**Supplementary Figure 5c**). These results demonstrate that Mpox SHINE can be applied directly to minimally processed samples while maintaining accuracy, albeit with delayed kinetics compared to extracted workflows.

## Discussion

This study demonstrates that highly sensitive CRISPR-based pathogen detection can be designed, optimized, and field-validated within the timeline of an active outbreak. In the middle of the 2025 Sierra Leone mpox surge, we went from sequence access to in-country clinical evaluation in less than three months **(Supplementary Figure 6)**. In practice, the design cycle itself was much faster: assay targets were identified within days of genome release, lyophilized reagents were prepared and validated within weeks, and integration onto a portable isothermal device required only minimal adaptation. With streamlined coordination, these steps could be compressed even further into a matter of days to weeks, underscoring the feasibility of moving from pathogen sequence to functional, field-ready diagnostics on outbreak timescales.

Mpox SHINE achieved strong performance on both extracted and unextracted clinical samples. On 56 extracted samples, all positives and negatives were correctly classified, with all positives detected within 37 minutes and most within 15 minutes. The same chemistry translated to unextracted lesion swabs processed by chemical lysis, with complete agreement across 16 additional samples. Time to detection on unextracted samples averaged under 30 minutes, placing actionable results well within the window for clinical decision-making or public health triage.

Together, these results establish Mpox SHINE as a portable, rapid, and accurate diagnostic that works not only on purified DNA but also on minimally processed samples, a critical step toward true POC deployment.

These findings add to a growing body of work showing that CRISPR-based and other isothermal diagnostics can be moved out of central laboratories and into the field (*20–26*). Prior SHINE studies demonstrated equipment-free workflows for SARS-CoV-2 and influenza (*17*, *18*), but this work extends the approach into the setting of an ongoing epidemic in West Africa, with assays validated on-site in Sierra Leone. By showing that the same chemistry can deliver single-digit copy sensitivity, reproducible performance on a portable fluorescence device, and compatibility with unextracted samples, we take an important step toward closing the diagnostic gap that has hampered outbreak response in resource-limited settings.

A key enabler of this work was the DxHub platform, which brings together programmable isothermal incubation, dual-channel fluorescence detection, and an intuitive workflow in a compact, field-deployable device. Unlike traditional SHINE readouts that rely on lateral flow strips, the DxHub provides quantitative fluorescence signal monitoring in real time, allowing for automated thresholding, data export, and integration into digital health systems. The device’s modular design supports not only Mpox SHINE but also a wide range of isothermal assays, creating a flexible platform for point-of-care testing. By combining robust assay chemistry with a deployable instrument, this collaboration highlights the translational potential of industry–academic partnerships in bringing next-generation molecular diagnostics to the front lines.

Further work can refine assay performance and address sources of variability. Two negative samples each produced late, low-amplitude signals in one of three replicates, though these did not compromise assay specificity due to interpretation on the sample level via a 2 of 3 majority rule.

Future work should focus on finer-tuned fluorescence threshold adjustment or by reflex testing any replicate that produces atypical signal profiles. Either approach would maintain the observed accuracy while further reducing the risk of false positives. In addition, thresholds for unextracted samples were set retrospectively using matrix-specific controls, reflecting the challenges of on-site assay calibration. Prospective validation in larger cohorts and across diverse sample types will be essential to confirm robustness.

This study illustrates the transformative potential of rapid, programmable diagnostics in outbreak response. By compressing the pathway from assay design to in-country validation, and by demonstrating accurate detection directly from unextracted clinical samples, Mpox SHINE charts a path toward truly field-ready CRISPR diagnostics. With further refinement, it is realistic to envision a future where new assays can be designed, validated, and deployed in real time as pathogens emerge. This work represents an early step toward that future, showing how global readiness can be strengthened by uniting advances in molecular biology, computational design, and portable instrumentation into a rapid-response diagnostic pipeline.

## Materials and Methods

### Mpox SHINE Assay Designs

*Guide RNA design.* Mpox genomes were retrieved from NCBI (TaxID 10244) with sequence length ≥180,000 bp and collection date ≥2022-01-01 (downloaded 2025-02-07). From these, 50 random genomes each from Clade IIb lineage B.1 (target set), Clade Ia and Clade Ib (non-target sets) were chosen. ADAPT (Activity-informed Design with All-inclusive Patrolling of Targets) (*14*) was used to generate candidate crRNA designs by inputting the Mpox IIb-B.1 sequences as the target set. Consensus sequences of Mpox Ia and Ib were computed after alignment using MAFFT via Geneious software and used in ADAPT as non-targets to ensure specificity of crRNA candidates to Mpox IIb B.1. ADAPT crRNA designs were mapped against a larger subset of mpox IIb-B.1, Ia, and Ib sequences (also obtained from NCBI Virus) to select for maximal inclusion to Mpox IIb and minimal predicted detection of Mpox Ia or Ib. A crRNA was considered to detect a sequence if the crRNA mapped to the sequence with ≤ 2 mismatches for on-target sequences or ≤ 3 mismatches for off-target sequences. To validate the assay on the Sierra Leone mpox outbreak strain, the assay was mapped against the first 67 sequences obtained from the active outbreak (Pathoplexus). No mismatches were detected in any of these 67 sequences. The crRNA aligns from nucleotides 156,555 to 156,582 of the mpox genome (NC_063383).

*Primer design.* RPA forward and reverse primers were also designed using ADAPT. ADAPT was run on a multiple sequence alignment of Mpox viral genome sequences obtained from Kenema Government Hospital (KGH), with the following parameters: 35.0% to 65.0% guanine-cytosine (GC) content, max. 3 mismatches between the primer and target, full coverage in 98% of the genomes. The forward and reverse primers align to the mpox genome (NC_063383) from nucleotides 156,525 to 156,554 and 156,583 to 156,612, respectively with 0 mismatches detected in any of the input sequences. Forward RPA primers were ordered with and without a T7 promoter sequence (5’-GAAATTAATACGACTCACTATAGGG-3’) appended upstream. Both forward and reverse RPA primers and crRNAs were purchased from Integrated DNA Technologies. A synthetic DNA target sequence was ordered as a double stranded DNA gene fragment (dsDNA) from Twist Biosciences for use as a positive control and during assay development.

### Sequence alignment and percent identity analysis

Full-length reference genomes were obtained from NCBI RefSeq for Vaccinia (NC_006998.1), Cowpox (NC_003663.2), and Camelpox (AY009089.1), while clades Ia, Ib, IIa and IIb mpox genomes (including sequences from the 2025 Sierra Leone outbreak) were sourced from Pathoplexus (*16*). To assess conservation of the SHINE assay amplicon (nt 156,525 to 156,612 with respect to NC_063383) across mpox clades and other Orthopoxviruses, we performed multiple sequence alignment using MAFFT v7.490 (*27*, *28*) implemented in Geneious Prime (Biomatters Ltd.) with default parameters (gap opening penalty = 1.53; offset value = 0.123). Pairwise percent identity values were then calculated in Geneious as the proportion of identical nucleotides across the alignment length, with mismatches and gaps counted as differences. Amplicon absence from clade I (Ia/Ib) lineages was confirmed by the presence of characteristic deletions spanning this locus after alignment.

### Mpox SHINE master mix preparation

SHINE reactions were performed using the following conditions: 1× Lyo buffer (20 mM HEPES pH 8.0, 150 mM D-Mannitol and 5 % w/v Sucrose), 45 nM LwaCas13a protein (Genscript, Z03486-100) resuspended in 1× storage buffer (50 mM Tris-HCl pH 7.5, 600 mM NaCl, 2 mM DTT, and 5% Glycerol), 62.5 nM 6-U quenched FAM reporter (IDT), 2 mM of each ribonucleoside triphosphate (rNTP) (NEB, N0466S), 1 U/μL murine RNase inhibitor (NEB, M0314L), 1 U/μL NextGen T7 RNA polymerase (Biosearch Technologies, 30223-1), 0.1 U/μL RNase H (NEB, M0297L), 240-360 nM Forward RPA primer (120-180 nM Fwd primer with T7 promoter and 120-180 nM Forward primer without T7 promoter), 240-360 nM Reverse RPA primer (IDT), and 22.5 nM crRNA (IDT). TwistAmp basic kit RPA pellets (TwistDx, TABAS03KIT) were used at a ratio of 1 pellet per 100 μL of master mix. The final master mix was aliquoted to single-reaction volumes in individual 0.2 mL tubes, flash frozen in liquid nitrogen, and immediately set to lyophilize in a Labconco FreeZone 70040 4.5L Freeze Dryer overnight at-50 and a vacuum pressure of <0.4 mbar. The following day, freeze-dried Mpox SHINE pellets were either used immediately or stored at-20 for later use.

### Mpox SHINE plate reader fluorescence data collection

Reaction pellets were rehydrated with a resuspension buffer (RB at 1x: 60 mM KCl, 3.5% w/v PEG-8000, and 25 mM Magnesium Acetate), leaving volume space to add synthetic gene target or water as negative control up to 30 μL final volume.

Fluorescence kinetics were measured on a Biotek Cytation 5 plate reader (Agilent, USA) with excitation at 485 nm and emission at 520 nm at 38 every 20 s for up to 90 mins. Plate reader derived SHINE data was analyzed as follows: fluorescence signal at the 60 min time point, after baseline subtraction, was recorded for each reaction condition and a threshold was set at 10 standard deviations (SD) higher than the mean negative template control (NTC) endpoint relative fluorescence units (RFU).

### Mpox SHINE DxHub fluorescence data collection

For DxHub reactions, fluorescence signals were measured on a commercial DxHub device (DxLab Inc., Somerville, Massachusetts, United States; manufactured under contract by Axxin, Eaglemont, Australia) at 38 every 20 s for up to 60 mins. For controls and extracted clinical samples, the DxHub was operated at 50% of the device’s maximum FAM signal detection sensitivity; for unextracted samples, a 10% FAM sensitivity was used. DxHub was powered using an AC/DC power adapter provided with the device or a portable power bank.

### Analytical LOD determination

A fixed fluorescence threshold was set as the mean plus 10 standard deviations of negative template controls (NTCs) after baseline subtraction. Reactions were scored as positive if the fluorescence crossed this threshold by 60 minutes. The LOD was defined as the lowest input concentration at which ≥95% of replicates were detected, or if not achieved, as the concentration range spanning the lowest copy number input with incomplete detection and the subsequently higher copy number input with complete detection.

### Production and transport of Mpox SHINE pellets for clinical sample testing in Sierra Leone

SHINE reaction pellets were prepared at the Broad Institute, lyophilized overnight (LabConco Freezone 4.5L freeze dryer), and packed in sets of 24 pellets in aluminum vacuum sealed pack to minimize fluorescence quenching and with a silica gel desiccant included inside to minimize moisture buildup. Twenty-five vacuum packs of over 500 individual lyophilized SHINE pellets were prepared for transit to Kenema, Sierra Leone. All cold items were packed in a Credo DURACUBE Reusable Cold Chain Shipper, which maintained a steady temperature of-20C during travel.

### Clinical samples tested

Clinical samples were collected in the form of skin-lesion swabs stored in viral transport media (VTM), with clinical negatives representing samples from expected close contacts. While ideally negative controls would include specimens from patients with other skin-lesion–causing illnesses or infections with related orthopox viruses, such samples were not available during the study period. A total of 57 clinical samples were provided for our study, with which we performed repeat reference qPCR. Of these, 46 samples resulted in cycle threshold (CT) values which the Radifast mpox qPCR kit protocol considered positive (CT ≤ 40) whereas 11 samples had undefined CTs and were therefore considered confirmed negatives. We excluded one sample with the Ct of 40 from the positive set due to the borderline reliability, resulting in a final positive sample set size of 45.

### Clinical sample extractions and RADI FAST qPCR

Clinical samples were extracted by lab personnel at the KGH molecular laboratory in a BSL-3 facility, using the QIAamp DNA Micro kit (Qiagen, 56304) using the protocol for Isolation of Genomic DNA from Tissues. Extractions were performed with an input volume of 400 μL of sample (Viral transfer media with swabbed lesion material) and an elution volume of 200 μL using water. Extracted DNA was aliquoted for different assays and stored at-20 until used. qPCR was performed on extracted clinical samples using the RADI FAST Mpox Detection Kit (IVD) (KH Medical, RV015), using all instructions provided by the kit manufacturer. Cycling conditions were as follows: hold at 95 for 20 s, cycle at 95 for 2 s and 60 for 5 s for 45 cycles. Real-time qPCR was run on a Roche LightCycler480. Data was analyzed using the LightCycler 480 II software version 1.5.1.62 SP3.

### Mpox SHINE testing for extracted clinical samples

Lyophilized SHINE pellets were stored at −20 °C; only the number required for a run (up to eight) were removed at a time. In a target-free, ventilated hood, each pellet was rehydrated with 15 µL of 2× Resuspension Buffer (120 mM KCl, 7.0% PEG-8000, 50 mM magnesium acetate), vortexed until fully dissolved, and briefly centrifuged. The resuspension was transferred to an individual dome-capped PCR tube. In a separate pre-amplification, target-handling hood, 15 µL of extracted clinical sample was added to the tube, mixed, and briefly spun down. Tubes were loaded into the DxHub and incubated at 38 °C and for a 60-min Mpox SHINE run, with fluorescence signals measured every 20 s. Each clinical sample was tested in three technical replicates, and at least 2–3 positive and 2–3 negative controls were included per test day.

### Assessment of compatibility of lysis buffers with mpox SHINE reaction

Initial testing was done on the plate reader to determine mpox SHINE reaction compatibility with the expected sample matrix (VTM) in combination with two different lysis buffers. Buffer 1, aka “Saliva Lysis Buffer”: 12.5 mM TCEP, 5 mM EDTA, 1% Pluronic, 55 mM NaOH (*29*); Buffer 2: 5x Fast-Amp Viral and Cell Lysis Solution (IntactGenomics, 4631)) was combined with VTM and included in mpox SHINE reactions at different volume proportions to determine effect on sensitivity and reaction kinetics **(Supplementary Figure 3a-c)**. Buffer 1 was chosen based on the observed reaction performance and tested on the DxHub at 30% volume **(Supplementary Figure 3d, Figure 3b-left side)**.

### Chemical lysis of unextracted mpox samples

Prior to testing of unextracted clinical samples, Buffer 1 was prepared in a target-free hood and aliquoted into PCR tubes (6 µL per tube; one tube per sample). Tubes were transferred into the BSL-3 laboratory, where 24 µL of unextracted clinical sample was added to each tube. After pipette mixing and a brief centrifugation, tubes were incubated at room temperature for ≥5 minutes to achieve chemical lysis and viral inactivation. The lysed samples were then ready for addition to SHINE reactions.

### Mpox SHINE testing for unextracted clinical samples

To minimize any potential autofluorescence and inhibitory interactions from unextracted matrices, pellets were rehydrated with a magnesium-only buffer (no PEG/KCl), formulated so that after sample addition the final Mg² concentration was 30 mM. In a target-free hood, each pellet was rehydrated with 21 µL, vortexed to fully dissolve, and transferred to a PCR tube. Tubes were then taken into the BSL-3 laboratory, where 9 µL of chemically lysed clinical sample was added, mixed, and briefly centrifuged. The assembled reactions were returned to the main lab and run on the DxHub. Because VTM raised baseline fluorescence on the device, the DxHub FAM sensitivity was set to 10% to prevent early saturation and improve interpretation.

### Fluorescence SHINE data analysis

Fluorescence data from both plate reader–derived and DxHub–derived SHINE assays were analyzed using a uniform approach. Fluorescence signal at the 60-min time point, after baseline subtraction (average RFU from 1–4 min), was recorded for each reaction condition. A positivity threshold was set at the mean endpoint fluorescence of negative template control (NTC) reactions plus 10 standard deviations. This conservative cutoff was selected to minimize the probability of false-positive calls, accounting for baseline variability and instrument noise across platforms. While lower multipliers (e.g., 3–5 SD) have been reported for similar isothermal and CRISPR-based assays, we applied a 10 SD cutoff to maximize robustness and reproducibility, particularly in the context of field-deployment where reaction matrices and operating conditions may vary.

### Statistical methods and sensitivity and specificity determination

Each clinical sample was tested in triplicate to provide statistical rigor, mitigate stochastic effects at low template concentrations, and enable application of replicate-based interpretation rules. At the sample level, we called results called positive if at least 2 of 3 replicates exceeded the threshold within the 60-min run window. For unextracted clinical samples, where duplicate reactions were run, we called a result positive if both (2 of 2) replicates exceeded the threshold.

Sensitivity (true positive rate) was defined as the proportion of qPCR-positive samples correctly identified as positive by SHINE, and specificity (true negative rate) as the proportion of qPCR-negative samples correctly identified as negative. Point estimates of sensitivity and specificity were calculated in GraphPad Prism (v10.2) with 95% one-sided confidence intervals using the Wilson score method, appropriate for small sample sizes and binomial outcomes.

All plots and statistical summaries were generated in GraphPad Prism, and results were confirmed by manual cross-checking of replicate data.

## Data Availability

All data supporting the findings of this study are available in the main text or the Supplementary Materials. Raw fluorescence and qPCR data underlying the figures are available from the corresponding authors upon reasonable request. Genome sequence data used for assay design are publicly available through Pathoplexus

## Acknowledgments

We thank Kenema Government Hospital (KGH) and Lassa Lab for samples, expertise, and invaluable support during the study.

The authors acknowledge the use of OpenAI’s ChatGPT (GPT-5, October 2025 version) to assist in improving the clarity and structure of the manuscript text. The tool was not used to generate or analyze data, create figures, or draw scientific conclusions. All content was reviewed, verified, and approved by the authors.

## Author contributions

Study design and conceptualization: NG, JDS

Mpox SHINE development: NG, CP, MG, TA

Clinical samples and extractions: JDS, DG, IUF, MMR, TPK, MSK

Clinical sample data collection and qPCR: NG, TA, IUF, MMR

DxHub integration: NG, HS

Writing and figures—original draft: NG

Writing and figures—review & editing: NG, PCS, AO, HS, LAB, DJR, JDS, TA, ES

Supervision: PCS, AO

## Ethical approvals

All research activities complied with relevant ethical regulations and institutional policies. Activities at Kenema Government Hospital (KGH) were conducted under approval from the Sierra Leone Ethics and Scientific Review Committee (protocol 002/05/2024). Activities at the Broad Institute were conducted under approvals from the Harvard Longwood Campus Institutional Review Board (protocols IRB24-0562, IRB24-0563).

## Data and materials availability

All data supporting the findings of this study are available in the main text or the Supplementary Materials. Raw fluorescence and qPCR data underlying the figures are available from the corresponding authors upon reasonable request. Genome sequence data used for assay design are publicly available through Pathoplexus (“Pathoplexus | Mpox Virus Genomes,” n.d.).

## Supplementary Materials

**Supplementary Figure S1.**
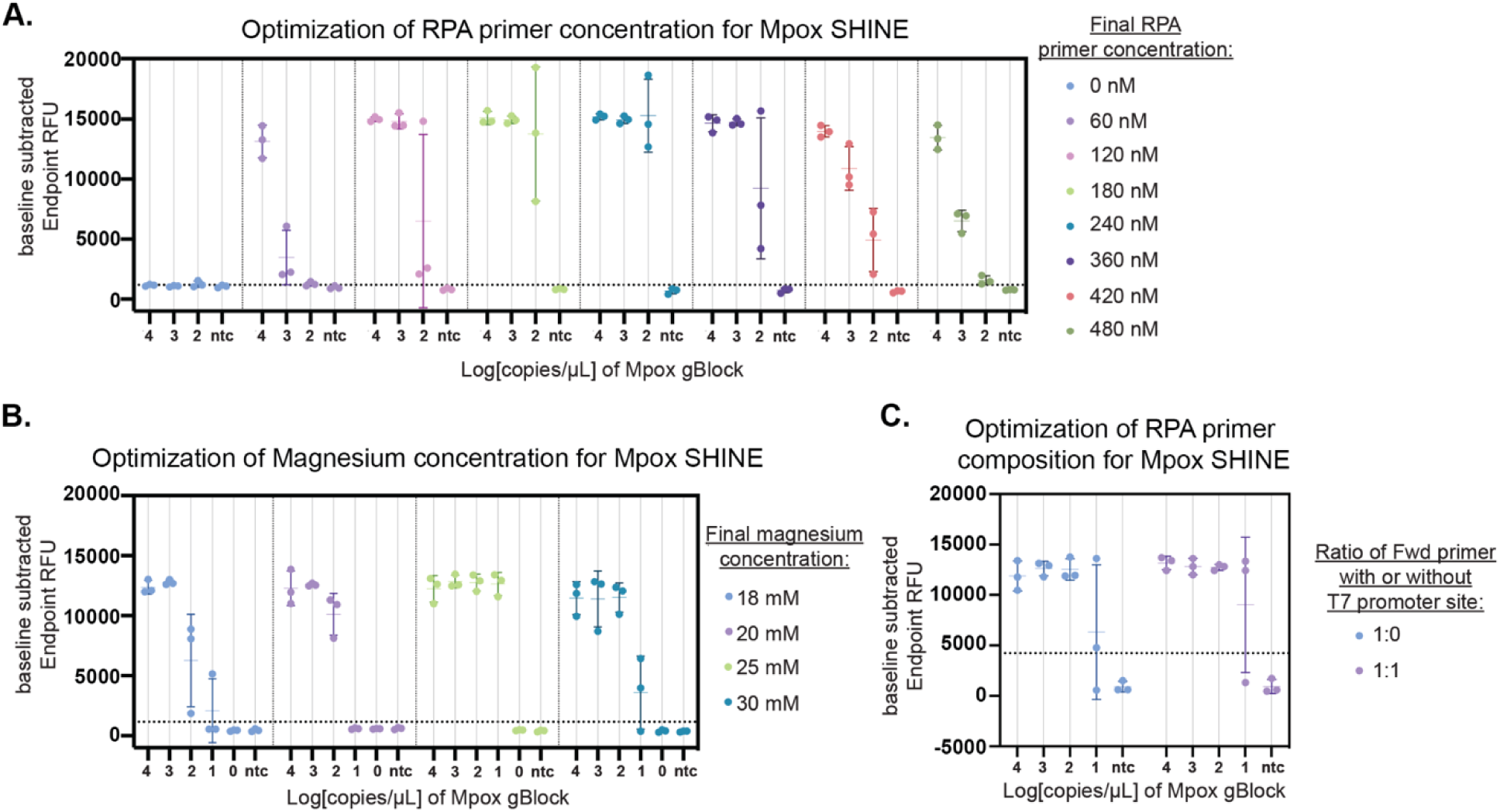
— Mpox SHINE optimization on plate reader. **(A)** Primer concentration titration. Forward and reverse RPA primers tested at the indicated final concentrations (0–480 nM each) against a log dilution of synthetic target (log4 – log0 copies/µL in the reaction) and no-template controls (NTC). 240 nM or 360 nM primer was used for subsequent experiments. **(B)** Magnesium titration. Final Mg² (magnesium acetate) was varied (18–30 mM) under the same target series. 25 mM Mg^2+^ was used for all subsequent experiments. **(C)** Forward-primer composition. Comparison of a 1:0 mix (100% primer with T7 promoter) versus a 1:1 mix (50% primer with T7 promoter and 50% primer without T7 promoter); reverse primer and other components held constant. A 1:1 mix was used in all subsequent experiments. For all panels, points are individual replicates and error bars show mean ± SD **of** endpoint fluorescence (baseline-subtracted RFU at 60 min; baseline = mean RFU from 1–4 min). The horizontal dashed line marks the fixed RFU calling threshold used for figure panels. Reactions were run at 38 °C. NTCs remained below threshold across conditions.

**Supplementary Figure 2.**
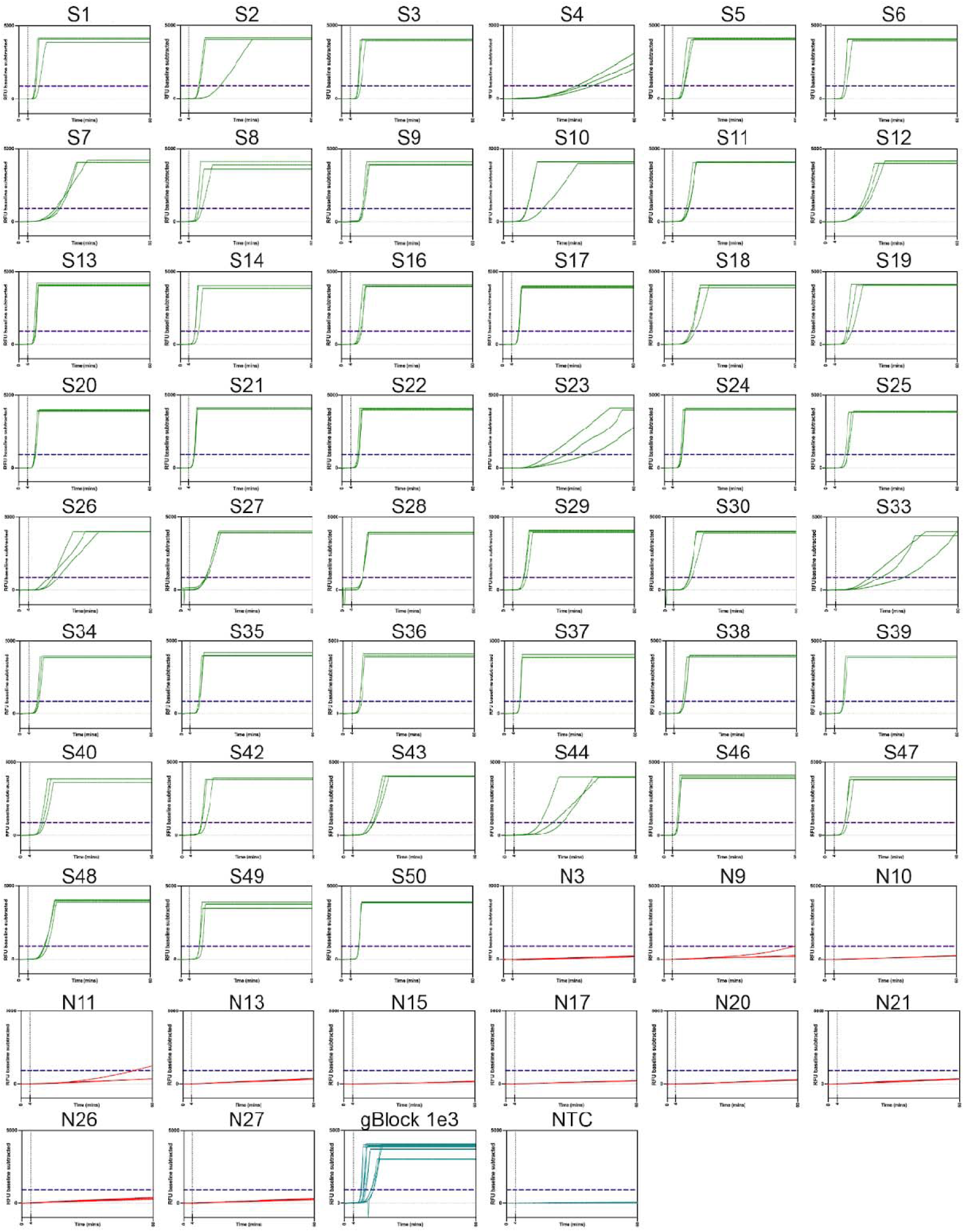
Individual fluorescence vs time traces for Mpox SHINE extracted **clinical sample tests.** Baseline-subtracted FAM fluorescence over time for individual cli ical specimens run in triplicate at 38 °C (one panel per specimen). Green traces = qPCR-positive samples; red traces = qPCR-negative samples. The horizontal dashed line marks the fixed RFU threshold. Last two panels in the final row include positive controls (10^3^ copies/µL gBlock) and no-template controls (NTCs).

**Supplementary Figure 3.**
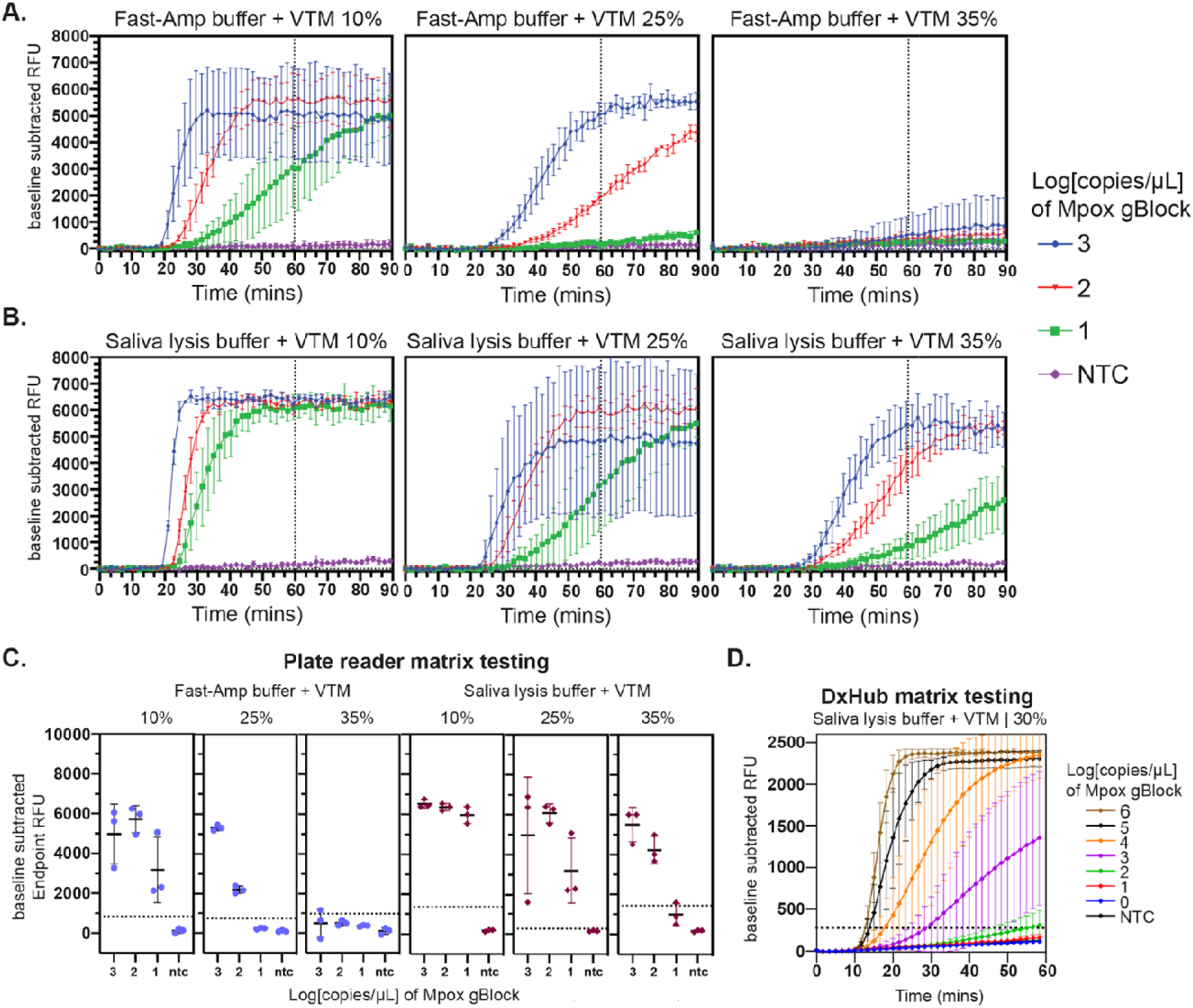
Mpox SHINE matrix testing on the plate reader. **(A-B)** Real-time FAM fluorescence (baseline-subtracted RFU) for contrived Mpox gBlock at three input levels (10^3^, 10^2^, 10^1^ copies/µL in the reaction; blue, red, green) and NTCs (magenta) in viral transport medium (VTM) mixed with either FastAmp lysis buffer (IntactGenomics, cat# 4631) **(A)** or Saliva Lysis Buffer (Li et al., 2023) **(B).** Each panel shows three final matrix fractions per reaction (10%, 25%, 35% VTM+lysis buffer). Lines/points denote the mean; error bars indicate SD across technical replicates. The vertical dashed line marks the **60-min** decision time. N = 3 technical replicates per condition. **(C)** Endpoint fluorescence (baseline-subtracted RFU) from the same experiments shown in panels **A** and **B**, summarized by condition (mean ± SD). Horizontal lines denote thresholds calculated per condition as NTC mean + 10 SD. **(D)** Real-time FAM fluorescence measured on the DxHub (baseline-subtracted RFU) for contrived Mpox gBlock at six input levels (10^6^ to 10^0^ copies/µL in the reaction) and NTCs in viral transport medium (VTM) mixed with saliva lysis buffer, included in the reaction at 30% volume (9 µL matrix per 30 µL reaction). DxHub FAM sensitivity set to 10% for reactions with matrix.

**Supplementary Figure 4.**
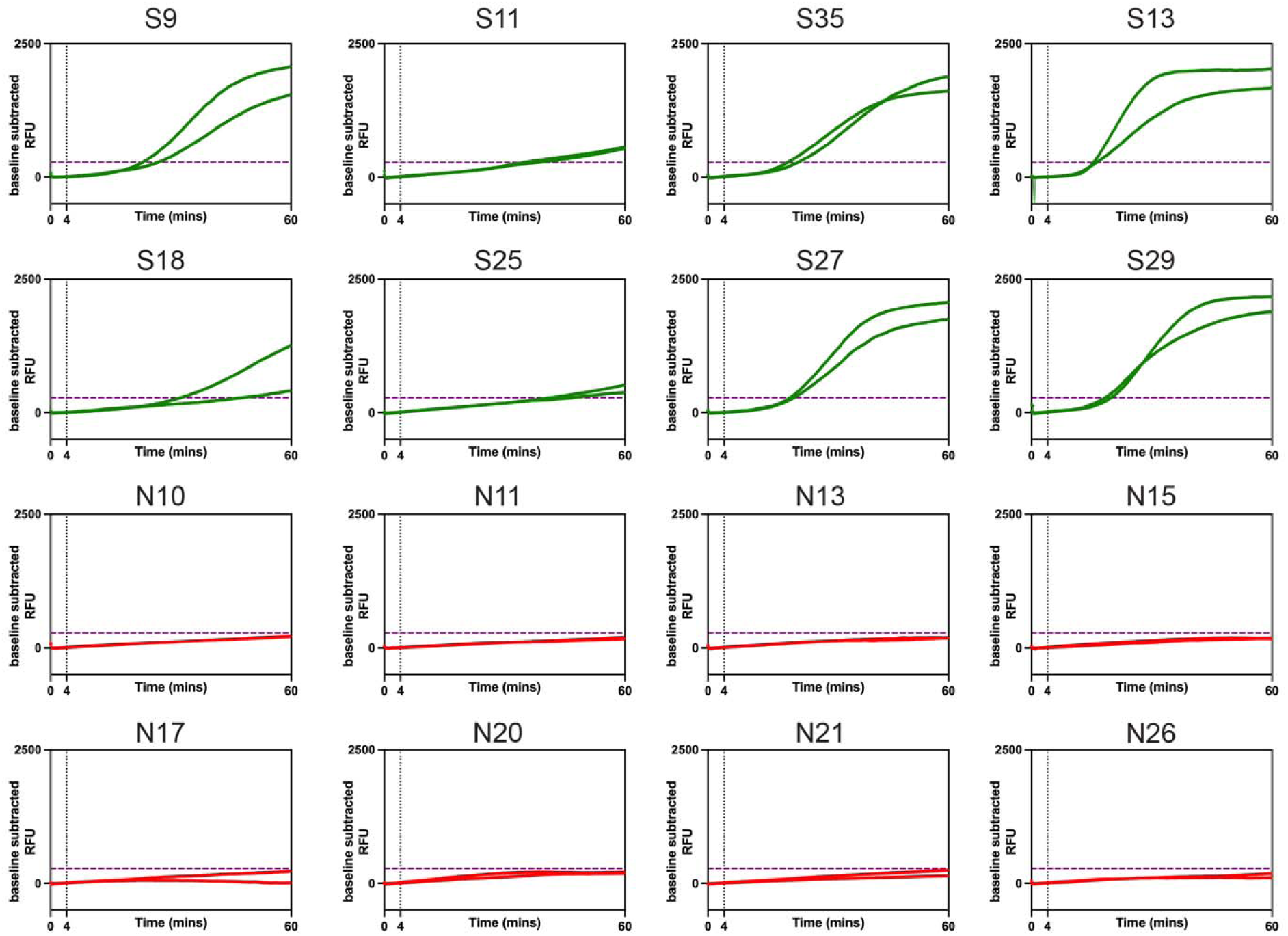
Individual fluorescence vs time traces for Mpox SHINE unextracted **clinical sample tests.** Baseline-subtracted FAM fluorescence over time for individual cli ical specimens run in triplicate at 38 °C (one panel per specimen). Green traces = qPCR-positive samples; red traces = qPCR-negative samples. The horizontal dashed line marks the fixed RFU threshold.

**Supplementary Figure 5.**
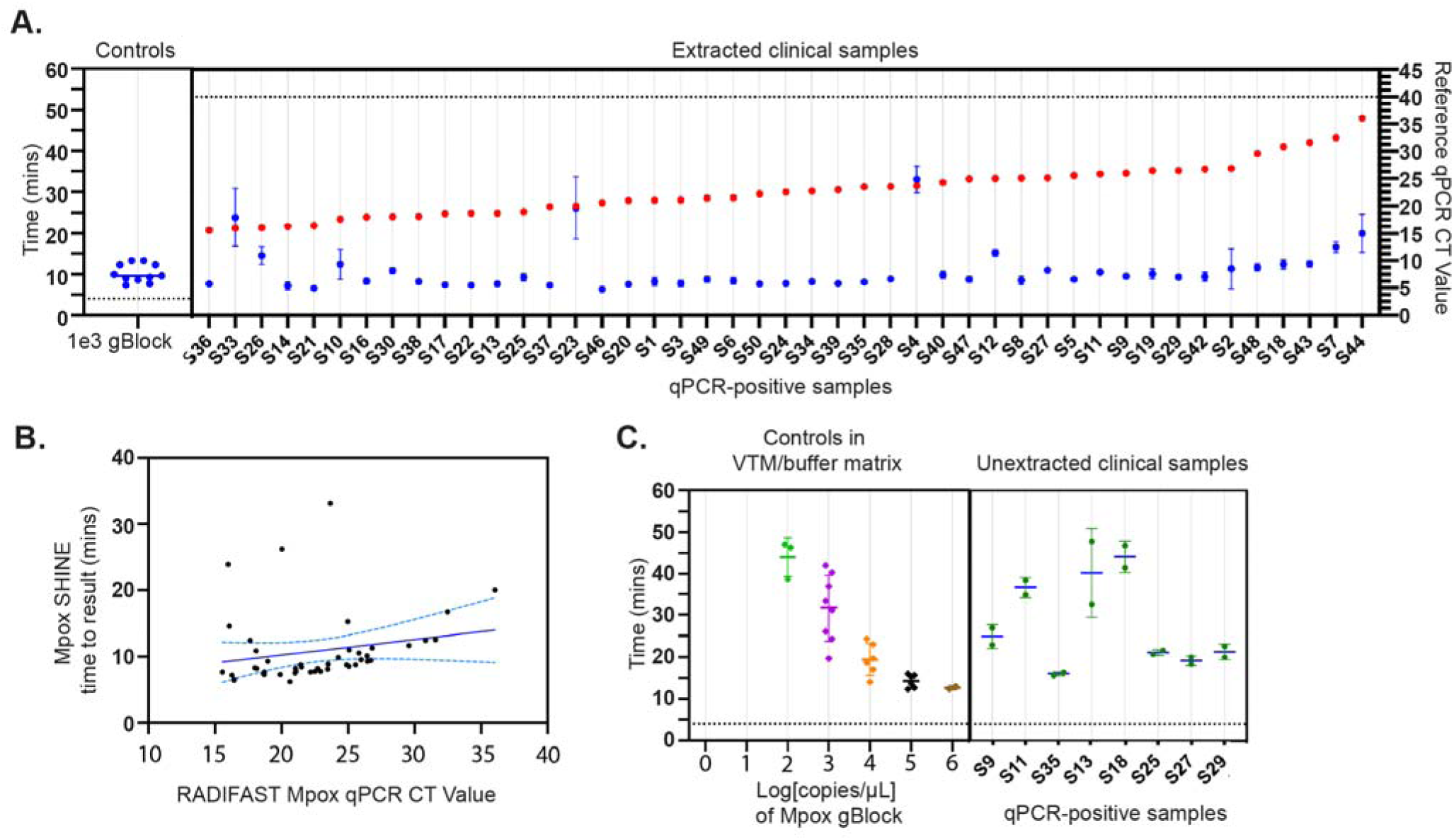
Relationship between qPCR Ct values and SHINE time-to-detection. **(A)** Time to detection (left y-axis) for controls and Qiagen-extracted clinical samples is plotted in minutes. Control samples (Mpox gBlock at 1e3 copies/µL, blue symbols, n=11) are shown on the left. qPCR-positive clinical samples are shown on the right (blue circles, n=47 positive samples), and the corresponding qPCR Ct values for each sample are shown as red circles (right y-axis). For each clinical sample, the mean ± SD across 3 technical replicates is plotted, and samples in this figure are plotted in CT value order (right y-axis) from smallest to largest left to right. **(B)** Scatterplot of Mpox SHINE time to result values versus corresponding RADIFAST qPCR CT values for extracted clinical samples (n=46). Solid line shows linear regression fit (R squared = 0.0412) with 95% confidence bands (dashed lines). Pearson correlation: Pearson r = 0.20, 95% CI −0.10–0.47, p = 0.1810, not significant. Spearman correlation: ρ = 0.46, 95% CI 0.18–0.67, p = 0.0016, mildly significant. **(C)** Time to result values for controls and unextracted clinical samples. Control reactions in VTM + saliva lysis buffer matrix is shown on the left with a 10-fold dilution series of Mpox gBlock (10^6^–10^0^ copies/µL of reaction; n = 2–8 per dilution (see **Figure 3b** for replicate details)). qPCR-positive unextracted clinical samples are shown on the right (blue circles, n=8 positive samples). For each clinical sample, the mean ± SD across 2 technical replicates is shown.

**Supplementary Figure 6.**
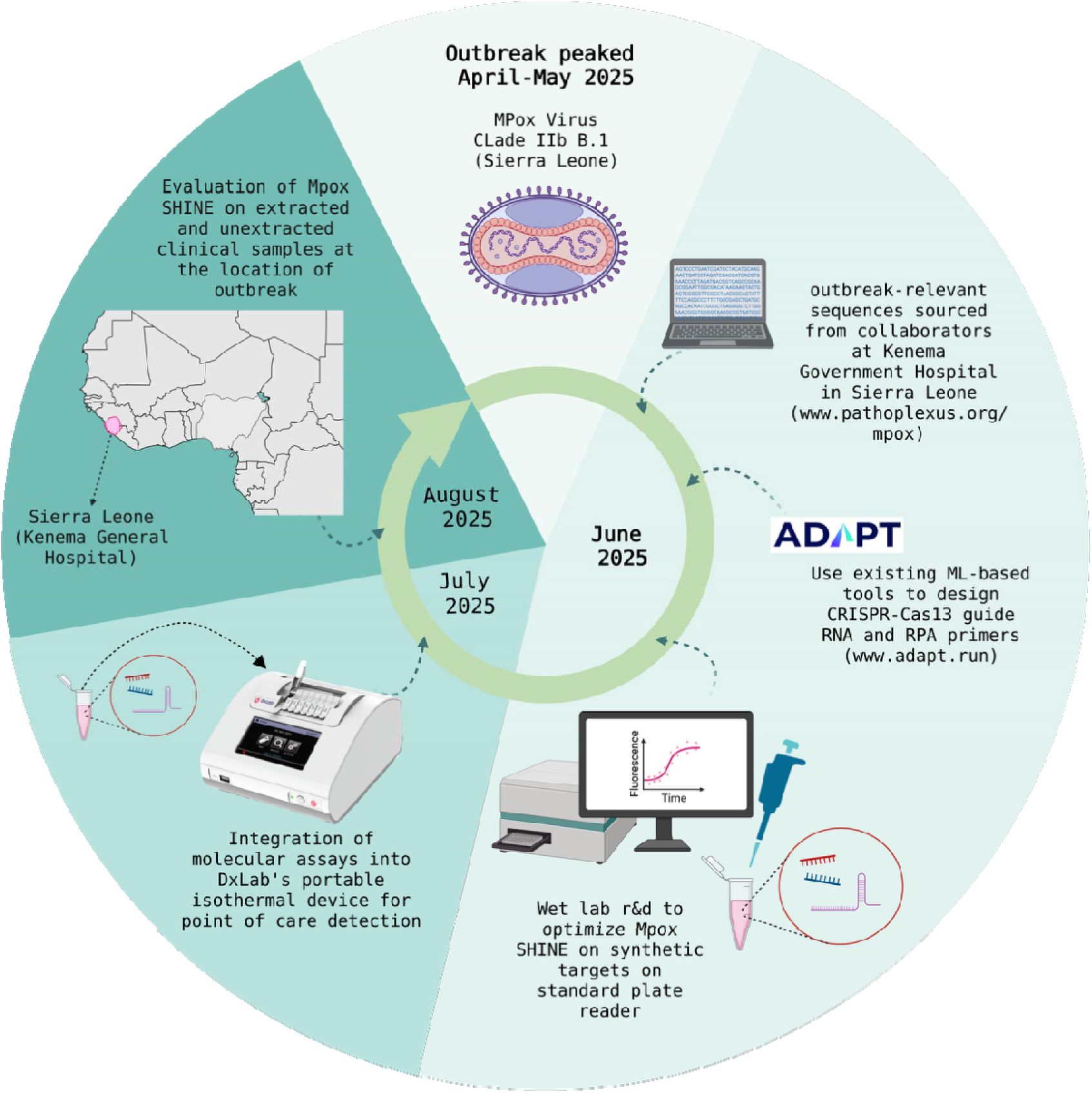
Timeline and workflow of assay design, development, device-integration, and in-country validation on clinical samples, in response to the Mpox Sierra L one 2025 outbreak. Figure made in Biorender.

---

**Supplementary Table 1.**
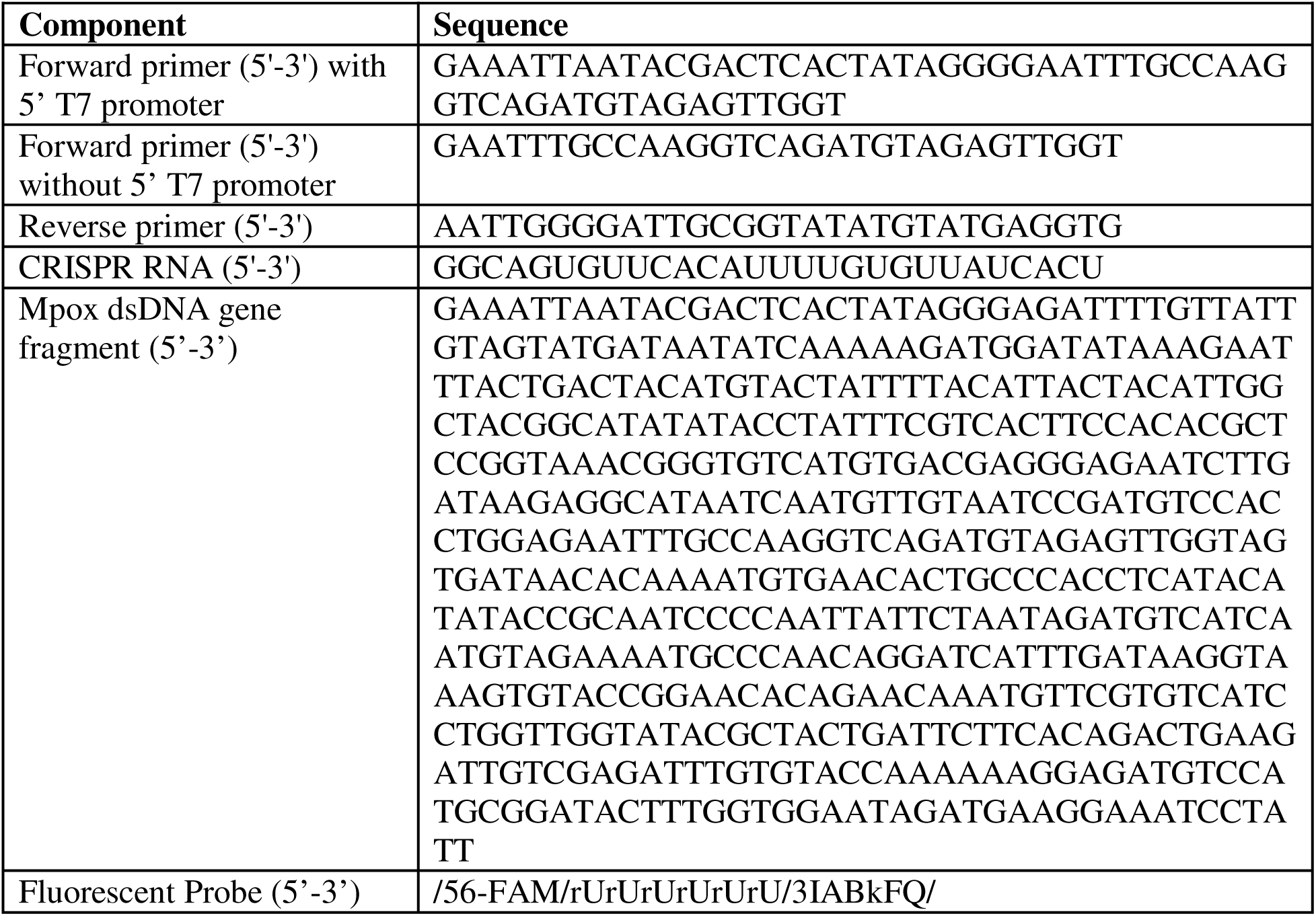
Assay designs.

**Supplementary Table 2.** Clinical Sample Cohort.

| Sample ID | KGH Sample ID | Mpox Clade IIb - qPCR CT | Internal Positive Control - qPCR CT | Positive SHINE result (replicates / 3) | Sample level SHINE result (majority rule) |
| --- | --- | --- | --- | --- | --- |
| S1 | G-29281-1 | 21.04 | 24.00 | 3 | Positive |
| S2 | G-29251-1 | 26.81 | 30.25 | 3 | Positive |
| S3 | G-29288-1 | 21.08 | 24.88 | 3 | Positive |
| S4 | G-29207-1 | 23.66 | 20.59 | 3 | Positive |
| S5 | G-29212-1 | 25.55 | 28.24 | 3 | Positive |
| S6 | G-29206-1 | 21.49 | 32.82 | 3 | Positive |
| S7 | G-29230-1 | 32.47 | 34.29 | 3 | Positive |
| S8 | G-29248-1 | 25.06 | 28.11 | 3 | Positive |
| S9 | G-29283-1 | 25.97 | 26.54 | 3 | Positive |
| S10 | G-29250-1 | 17.63 | 19.57 | 3 | Positive |
| S11 | G-29223-1 | 25.80 | 24.29 | 3 | Positive |
| S12 | G-29269-1 | 24.98 | 27.99 | 3 | Positive |
| S13 | G-29272-1 | 18.70 | 21.78 | 3 | Positive |
| S14 | G-29291-1 | 16.28 | 19.55 | 3 | Positive |
| S16 | G-29222-1 | 17.99 | 20.10 | 3 | Positive |
| S17 | G-29205-1 | 18.63 | 21.80 | 3 | Positive |
| S18 | G-29193-1 | 30.82 | 34.87 | 3 | Positive |
| S19 | G-29247-1 | 26.43 | 29.01 | 3 | Positive |
| S20 | G-29218-1 | 21.01 | 24.24 | 3 | Positive |
| S21 | G-29241-1 | 16.45 | 19.75 | 3 | Positive |
| S22 | G-29217-1 | 18.69 | 22.03 | 3 | Positive |
| S23 | G-29204-1 | 20.02 | 21.10 | 3 | Positive |
| S24 | G-29236-1 | 22.51 | 24.69 | 3 | Positive |
| S25 | G-29216-1 | 18.96 | 22.48 | 3 | Positive |
| S26 | G-29203-1 | 16.07 | 18.48 | 3 | Positive |
| S27 | G-29268-1 | 25.07 | 27.83 | 3 | Positive |
| S28 | G-29239-1 | 23.49 | 26.54 | 3 | Positive |
| S29 | G-29215-1 | 26.45 | 29.60 | 3 | Positive |
| S30 | G-29202-1 | 18.08 | 20.80 | 3 | Positive |
| S33 | G-29264-1 | 15.99 | 18.27 | 3 | Positive |
| S34 | G-29235-1 | 22.71 | 25.33 | 3 | Positive |
| S35 | G-29260-1 | 23.45 | 26.01 | 3 | Positive |
| S36 | G-29234-1 | 15.56 | 18.88 | 3 | Positive |
| S37 | G-29213-1 | 19.90 | 23.36 | 3 | Positive |
| S38 | G-29200-1 | 18.12 | 21.58 | 3 | Positive |
| S39 | G-29210-1 | 22.93 | 24.99 | 3 | Positive |
| S40 | G-29199-1 | 24.25 | 25.29 | 3 | Positive |
| S42 | G-29233-1 | 26.70 | 29.06 | 3 | Positive |
| S43 | G-29257-1 | 31.58 | 34.28 | 3 | Positive |
| S44 | G-29231-1 | 36.04 | 38.28 | 3 | Positive |
| S46 | G-29232-1 | 20.62 | 24.65 | 3 | Positive |
| S47 | G-29209-1 | 24.89 | 28.08 | 3 | Positive |
| S48 | G-29208-1 | 29.57 | 32.15 | 3 | Positive |
| S49 | G-29196-1 | 21.42 | 24.81 | 3 | Positive |
| S50 | G-29197-1 | 22.17 | 25.48 | 3 | Positive |
| N3 | 29275 |  | 32.70 | 0 | Negative |
| N9 | 29046 |  | 23.30 | 1 | Negative |
| N10 | 29045 |  | 27.85 | 0 | Negative |
| N11 | 29073 |  | 27.41 | 1 | Negative |
| N13 | 29068 |  | 30.68 | 0 | Negative |
| N15 | 29018 |  | 20.87 | 0 | Negative |
| N17 | 29274 |  | 21.92 | 0 | Negative |
| N20 | 29287 |  | 27.65 | 0 | Negative |
| N21 | 29286 |  | 22.45 | 0 | Negative |
| N26 | 29279 |  | 30.24 | 0 | Negative |
| N27 | 29278 |  | 22.60 | 0 | Negative |

**Supplementary Table 3.** Mpox clade IIb accession numbers via Pathoplexus.

| Pathoplexus<br>Accession | District in<br>Sierra<br>Leone | length | Submissi<br>on ID | Release Date | completeness | Total<br>Ambiguous<br>Nucs | Total<br>Unknown<br>Nucs |
| --- | --- | --- | --- | --- | --- | --- | --- |
| PP_002XLG<br>K.1 | Kenema | 197184 | G-29022-1 | 5/27/25 | 0.99997972 | 3 | 1 |
| PP_002XLH<br>G.1 | Kenema | 197112 | G-29023-1 | 5/27/25 | 0.99969068 | 3 | 58 |
| PP_002XKV<br>T.1 | Kailahun | 197106 | G-28995-1 | 5/27/25 | 0.99998479 | 3 | 0 |
| PP_002XKU<br>V.1 | Kenema | 197190 | G-28994-1 | 5/27/25 | 0.99998479 | 3 | 0 |
| PP_002XL0H<br>.1 | Bo | 197121 | G-29004-1 | 5/27/25 | 0.99997972 | 4 | 0 |
| PP_002XKW<br>R.1 | Kenema | 197156 | G-29000-1 | 5/27/25 | 0.99997465 | 5 | 0 |
| PP_002XKX<br>P.1 | Bo | 197157 | G-29001-1 | 5/27/25 | 0.99997972 | 4 | 0 |
| PP_002XKSZ<br>.1 | Kenema | 197183 | G-28997-1 | 5/27/25 | 0.99997972 | 4 | 0 |
| PP_002XKT<br>X.1 | Kenema | 197169 | G-28998-1 | 5/27/25 | 0.99927995 | 9 | 125 |
| PP_002XLN6<br>.1 | Kenema | 197080 | G-28999-1 | 5/27/25 | 0.99947771 | 3 | 92 |
| PP_002XKY<br>M.1 | Bo | 197149 | G-29002-1 | 5/27/25 | 0.99997465 | 5 | 0 |
| PP_002XKZ<br>K.1 | Bo | 197140 | G-29003-1 | 5/27/25 | 0.99998479 | 3 | 0 |
| PP_002XL1F<br>.1 | Bo | 197180 | G-29005-1 | 5/27/25 | 0.99998479 | 3 | 0 |
| PP_002XL2D<br>.1 | Bonthe | 197138 | G-29007-1 | 5/27/25 | 0.99997465 | 5 | 0 |
| PP_002XLB<br>V.1 | Kenema | 197178 | G-29008-1 | 5/27/25 | 0.99956899 | 5 | 80 |
| PP_002XL49.<br>1 | Bo | 197186 | G-29011-1 | 5/27/25 | 0.99993915 | 3 | 9 |

| Pathoplexus Accession | District in Sierra Leone | length | Submission ID | Release Date | completeness | Total Ambiguous Nucs | Total Unknown Nucs |
| --- | --- | --- | --- | --- | --- | --- | --- |
| PP_002XL57.1 | Bo | 197179 | G-29012-1 | 5/27/25 | 0.99990873 | 11 | 7 |
| PP_002XL65.1 | Bo | 197142 | G-29013-1 | 5/27/25 | 0.99996958 | 6 | 0 |
| PP_002XL3B.1 | Kenema | 197173 | G-29010-1 | 5/27/25 | 0.99997465 | 5 | 0 |
| PP_002XL73.1 | Kenema | 197175 | G-29014-1 | 5/27/25 | 0.9999645 | 7 | 0 |
| PP_002XLD R.1 | Bonthe | 197170 | G-29015-1 | 5/27/25 | 0.99982759 | 3 | 31 |
| PP_002XLEP.1 | Bonthe | 197155 | G-29016-1 | 5/27/25 | 0.99997972 | 4 | 0 |
| PP_002XLF M.1 | Bonthe | 197171 | G-29017-1 | 5/27/25 | 0.99946757 | 4 | 101 |
| PP_002XL81.1 | Bonthe | 197153 | G-29019-1 | 5/27/25 | 0.99998479 | 3 | 0 |
| PP_002XL9Z.1 | Bonthe | 197119 | G-29020-1 | 5/27/25 | 0.99998479 | 3 | 0 |
| PP_002XLA X.1 | Kenema | 197093 | G-29021-1 | 5/27/25 | 0.99997972 | 4 | 0 |
| PP_002XLJE.1 | Kenema | 197177 | G-29025-1 | 5/27/25 | 0.99998479 | 3 | 0 |
| PP_002XLK C.1 | Kailahun | 197137 | G-29027-1 | 5/27/25 | 0.99997972 | 4 | 0 |
| PP_002XLL A.1 | Kailahun | 197132 | G-29028-1 | 5/27/25 | 0.99991887 | 9 | 7 |
| PP_002XLM 8.1 | Kailahun | 197163 | G-29029-1 | 5/27/25 | 0.99997972 | 4 | 0 |
| PP_0031UPN.1 | Kenema | 197180 | G-29030-1 | 7/8/25 | 0.99993915 | 0 | 12 |
| PP_0031UQL.1 | Bo | 197183 | G-29034-1 | 7/8/25 | 0.99967547 | 0 | 64 |
| PP_0031URJ.1 | Kenema | 197178 | G-29036-1 | 7/8/25 | 0.99700825 | 2 | 580 |
| PP_0031USG.1 | Kenema | 197178 | G-29038-1 | 7/8/25 | 0.99886415 | 0 | 216 |
| PP_0031UTE | Kenema | 197183 | G- | 7/8/25 | 0.99977182 | 1 | 44 |
| .1 |  |  | 29040-1 |  |  |  |  |
| PP_0031UUC<br>.1 | Kenema | 197172 | G-29041-1 | 7/8/25 | 0.99984281 | 0 | 23 |
| PP_0031UV<br>A.1 | Kenema | 197183 | G-29042-1 | 7/8/25 | 0.99995943 | 0 | 8 |
| PP_0031UW<br>8.1 | Kenema | 197176 | G-29043-1 | 7/8/25 | 0.99996958 | 0 | 0 |
| PP_0031UX6<br>.1 | Kenema | 197180 | G-29044-1 | 7/8/25 | 0.99997465 | 0 | 5 |
| PP_0031UY4<br>.1 | Kenema | 197183 | G-29049-1 | 7/8/25 | 0.99957406 | 0 | 84 |
| PP_0031UZ2.<br>1 | Kenema | 197183 | G-29050-1 | 7/8/25 | 0.99997465 | 1 | 4 |
| PP_0031V00.<br>1 | Bonthe | 197175 | G-29051-1 | 7/8/25 | 0.9999138 | 2 | 7 |
| PP_0031V1Y<br>.1 | Bonthe | 197180 | G-29052-1 | 7/8/25 | 1 | 0 | 0 |
| PP_0031V2<br>W.1 | Bonthe | 197175 | G-29053-1 | 7/8/25 | 0.99986309 | 1 | 18 |
| PP_0031V3U<br>.1 | Bonthe | 197157 | G-29054-1 | 7/8/25 | 0.99973125 | 0 | 53 |
| PP_0031V4S.<br>1 | Bonthe | 197174 | G-29058-1 | 7/8/25 | 0.99999493 | 1 | 0 |
| PP_0031V5Q<br>.1 | Kailahun | 197176 | G-29059-1 | 7/8/25 | 0.99973632 | 0 | 52 |
| PP_0031V7L.<br>1 | Bo | 197183 | G-29065-1 | 7/8/25 | 0.99998479 | 2 | 1 |
| PP_0031V8J.<br>1 | Bo | 197183 | G-29066-1 | 7/8/25 | 0.99996958 | 1 | 5 |
| PP_0031V6N<br>.1 | Pujehun | 197179 | G-29061-1 | 7/8/25 | 0.99995436 | 9 | 0 |
| PP_0031V9G<br>.1 | Kenema | 197191 | G-29070-1 | 7/8/25 | 0.99995943 | 0 | 0 |
| PP_0031VAE<br>.1 | Kenema | 197186 | G-29071-1 | 7/8/25 | 0.99661273 | 0 | 668 |
| PP_0031VD8<br>.1 | Kenema | 197186 | G-29110-1 | 7/8/25 | 0.99993915 | 0 | 12 |
| PP_0031VBC.1 | Kenema | 197176 | G-29102-1 | 7/8/25 | 0.99972111 | 0 | 55 |
| PP_0031VE6.1 | Kenema | 197199 | G-29111-1 | 7/8/25 | 0.99981745 | 1 | 35 |
| PP_002XLCT.1 | Kenema | 197136 | G-29009-1 | 5/27/25 | 0.99993408 | 3 | 2 |
| PP_0031VCA.1 | Kenema | 197184 | G-29106-1 | 7/8/25 | 0.99977182 | 1 | 44 |
| PP_0031VF4.1 | Bonthe | 197186 | G-29125-1 | 7/8/25 | 0.9991329 | 1 | 170 |
| PP_0031VG2.1 | Bonthe | 197180 | G-29126-1 | 7/8/25 | 0.99999493 | 1 | 0 |
| PP_0031VHZ.1 | Bonthe | 197179 | G-29127-1 | 7/8/25 | 0.99893514 | 1 | 201 |
| PP_0031VJX.1 | Bonthe | 197185 | G-29128-1 | 7/8/25 | 1 | 0 | 0 |
| PP_0031VK V.1 | Bonthe | 197176 | G-29129-1 | 7/8/25 | 1 | 0 | 0 |
|  |  |  | G-29006-1 |  |  |  |  |
|  |  |  | G-29024-1 |  |  |  |  |

